# Superantigenic TCR Vbeta 21.3 signature in Multisystem Inflammatory Syndrome in Children

**DOI:** 10.1101/2021.02.11.21251166

**Authors:** Marion Moreews, Kenz Le Gouge, Alicia Bellomo, Christophe Malcus, Rémi Pescarmona, Samira Khaldi-Plassart, Sophia Djebali, Anne-Laure Mathieu, Magali Perret, Marine Villard, Emilie Chopin, Isabelle Rouvet, Francois Vandenesh, Céline Dupieux, Robin Pouyau, Sonia Teyssedre, Margaux Guerder, Tiphaine Louazon, Anne-Moulin-Zinsch, Marie Duperril, Hugues Patural, Lisa Giovannini-Chami, Aurélie Portefaix, Behrouz Kassai, Fabienne Venet, Guillaume Monneret, Christine Lombard, Hugues Flodrops, Paul Bastard, Shen-Ying Zhang, Valérie Dubois, Olivier Thaunat, Jean-Christophe Richard, Mehdi Mezidi, Laurent Abel, Jean Laurent Casanova, Jacqueline Marvel, Sophie Trouillet-Assant, David Klatzmann, Thierry Walzer, Marlène Dreux, Encarnita Mariotti-Ferrandiz, Etienne Javouhey, Alexandre Belot

## Abstract

**Objectives:** Multiple Inflammatory Syndrome in Children (MIS-C) is the most severe pediatric form of COVID-19 and occurs in previously healthy children. MIS-C combines features of Kawasaki disease and Toxic Shock Syndrome (TSS).

**Methods:** Children with suspected MIS-C were included within the first week of diagnosis and a large scale immunoassay was performed to determein the immunologic signature of these patients.

**Results:** We characterized the immunological profile of 27 MIS-C cases in comparison with 4 KD and 4 TSS cases. Similarly to TSS, an increase of serum inflammatory cytokines (IL-6, TNF-a, CD25s) was observed in MIS-C contrasting with low expression of HLA-DR monocytes, a feature often associated with immune paralysis. Expansions of T cells expressing the Vβ21.3 T cell receptor β chain variable region were detected in both CD4 and CD8 subsets in almost 50% of patients and Vβ21.3-positive T cells expressed high level of HLA-DR highlighting their specific activation. TCR sequencing uncovered the polyclonal nature of the Vβ 21.3+ population. SARS-CoV2 antigene-specific production of interferon gamma in T cells was not increased in MIS-C T cells compared to COVID-19 patients suggesting the antigen-specific immune response in MIS-C patients is not pivotal to the manifestation.

**Conclusions:** Our findings argue in favor of a strong activation of the immune system related to a superantigenic immune response in MIS-C with a specific polyclonal Vβ21.3 T cell expansion.

**Key messages:** What is already known about this subject ?

MIS-C occurs 3-5 weeks after acute SARS-CoV2 infection and overlap features of Toxic Shock syndrome and Kawasaki disease.

MIS-C appears different in term of cytokine and autoantibodies generation from KD with subtle signs of T cells activation

What does this study add?

This study demonstrates that Vβ21.3+ CD4 and CD8 T cells are highly increased in about 50% of MIS-C and distinctive of the Vβ2+ expansion observed in toxic shock syndrome in This reflects a specific T cell activation and cytokine release syndrome similar to toxic shock syndrome

How mich this impact on clinical practice or future developments?

Vβ21.3+ signature can be available on a short term basis by flowcytometry and represents a signature of the MIS-C.

As for TSS, immunomodulating therapies may revert the superantigenic activation and resolve this life threatening pediatric condition.

## Introduction

At the end of April 2020, European clinicians warned the Public Health Agencies about an abnormal increase of Kawasaki-like diseases (KLD) and myocarditis requiring critical care support in the context of the ongoing COVID-19 epidemic in children(1–3). Later on, American clinicians also reported a large outbreak of severe inflammation in children following COVID-19 infection, a condition that is now named Pediatric Inflammatory Multisystemic Syndrome (PIMS) or Multisystem Inflammatory Syndrome in children (MIS-C)(4–6). The clinical phenotype of this emerging disease is broad and encompasses features of Kawasaki diseases (KD) and toxic shock syndrome (TSS). Many cases require intensive care support, making MIS-C one of the most severe manifestation of COVID-19 in children. Of note, the temporal distribution raises the hypothesis of a post-infectious disease occurring about 3 to 4 weeks after acute COVID-19 in children(3, 5–7).

For now, reports on MIS-C have shown slight differences in cytokine profiling and immunophenotype between MIS-C and KD or pediatric COVID-19(8, 9). Analysis of T cells revealed a lower number of T cells in MIS-C with no or subtle signs of activation(10). Multi-dimensional immune profiling on small numbers of patients showed differences between acute COVID-19 or pre-pandemic Kawasaki disease(8, 11). Anti-SARS-CoV2 antibodies were equally produced in pediatric COVID-19 and MIS-C. However, autoantibodies were uniquely produced during MIS-C or KD, which supports a contribution of the humoral response to both diseases(8, 11). Finally, a role for genetic factors has been evocated in MIS-C pathogenesis as it seems to occur more frequently in children from Hispanic or African ethnicity(12, 13).

Considering the similarities between MIS-C, KD and TSS, it has been hypothesized that SARS-CoV2 may act as a superantigen and induce a hyperinflammatory state in MIS-C(14, 15). Superantigens are molecular structures enabling polyclonal T cell response through binding external region of T cell receptor and MHC class II molecules(16). Consequently, superantigens can induce large expansions of T cells expressing one specific T-cell receptor (TCR) beta-chain variable region while classical antigens induce the expansion of T cells bearing different V□. For example, TSS is secondary to the effect of Staphylococcal TSS Toxin-1 (TSST-1) and leads to Vβ2+ T cell expansion(17). In KD, previous reports mentioned the expansion of different Vβ including Vβ2 in response to putative TSST1 but this point is controversial and there could be other causes and triggers of KD(14).

Here, we sought to test the hypothesis of a superantigen-type of T cell activation occurring in MIS-C. For this, we explored the cytokine and cellular immune profile in MIS-C and discovered a state of cytokine storm contrasting with a decrease in T cells and a reduction in their HLA-DR expression. We identified by flow cytometry a specific V□21;.3 signature in 12/25 tested patients with an expansion of more than twice the mean normal values, highlighting a superantigenic type of T cell response in MIS-C. TCR sequencing revealed the polyclonal nature of the Vβ21.3+ expansion. No specific HLA bias was identified in patients but we found a specific activation profile within Vβ21.3+ T cells.

## Methods

### Patients and ethics

Three distinct cohorts were used for the data collection and analyses registred in ClinicalTrial.gov and consent were obtained from parents. Details on IRB and ethical committee are available in supplemental data.

### Cytokines and IFN score assessment

Whole blood was sampled on EDTA tubes and plasma was frozen at −20°C within 4 hours following blood collection. Plasma concentrations of IL-6, TNF-a, IFN-g, IL-10, MCP-1, IL-1ra and CD25s were measured by Simpleplex technology using ELLA instrument (ProteinSimple), following manufacturer’s instructions. Plasma IFN-α concentrations were determined by single-molecule array (Simoa) on a HD-1 Analyzer (Quanterix) using a commercial kit for IFN-α2 quantification (Quanterix). Whole blood was collected on PAXgene blood RNA tubes (BD Biosciences) or on EDTA tubes for IFN signature, RNA extraction was performed with the kit maxwell 16 LEV simply RNA blood associated with the Maxwell extractor (Promega) and quantified by absorbance (Nanovue). IFN score was obtained using nCounter analysis technology (NanoString Technologies) by calculating the mediane of the normalized count of 6 ISGs as previously described(18)

### T-cell Vβ repertoire analysis and immunophenotyping

CD3, CD4 and CD8 T lymphocyte subsets were enumerated on EDTA-anticoagulated peripheral whole blood by single-platform the fully automated volumetric single plateforme technology flow cytometer AQUIOS CL (Beckman-Coulter) as previously described(19).

The phenotypic analysis of T-cell Vβ repertoire was performed on whole blood sample using the IOTest Beta Mark kit (Beckman-Coulter) containing 24 monoclonal antibodies (mAbs) identifying ∼ 70% of the T cell repertoire. Details are provided in supplemental data.

### Monocyte HLA-DR expression assessment

Monocyte HLA-DR expression was determined on EDTA-anticoagulated peripheral whole blood as previously describedde(20).

### TCR-sequencing

RNA was extracted from whole blood as reported above. T cell receptor (TCR) alpha/beta libraries were prepared from 300ng of RNA from each sample with SMARTer Human TCR a/b Profiling Kit (Takarabio) following provider protocol as previously described (32). Briefly, the reverse transcription was performed using a mixture of TRBC and TRAC reverse primers and further extended with a template-switching oligonucleotide (SMART-Seq v4). cDNAs were then amplified following two semi-nested PCR: a first PCR with TRBC and TRAC reverse primers as well as a forward primer hybridizing to the SMART-Seq v4 sequence added by template-switching and a second PCR targeting the PCR1 amplicons with reverse and forward primer including Illumina Indexes allowing for sample barcoding. PCR2 are then purified using AMPUre XP beads (Beckman-Coulter). The sequencing was then carried out on a NOVAseq 6000 Illumina sequencer using 2×250bp read length protocol. Paired-end sequences were aligned and annotated using MiXCR 3.0.13(21)

### Repertoire analysis

Analysis were performed in R 4.0.3 on the clonotype lists obtained with MiXCR. For each clonotype, read count was recorded. Frequencies for TRBV, TRBJ and clonotypes were calculated based on the total read counts per sample. Chord diagrams were made using the circlize package(22) on TRBVBJ frequencies, CDR3 length barplots were made using ggplot2 (23) on clonotype frequencies.

### Stimulation with SARS-CoV-2 overlapping peptide pools and flow cytometry

Briefly, overnight-rested PBMCs were stimulated with SARS-CoV-2 PepTivator pooled peptides (Miltenyi Biotec) at a final concentration of 2□μg□ml−1 for 1□h in the presence of 2□μg□ml−1 monoclonal antibodies CD28 and CD49d, and then for an additional 5□h with GolgiPlug and GolgiStop (BD Biosciences). Dead cells were labeled using LIVE/DEAD Fixable eF780 dye from Invitrogen. Surface markers, including APC-conjugated anti-CD3, BUV486-conjugated anti-CD4, PE-Cy7-conjugated anti-CD8, APC-Cy7-conjugated anti-CD14, APC-Cy7-conjugated anti-CD16 and APC-Cy7-conjugated anti-CD19 (BioLegend) were stained. Cells were then washed, fixed with Cytofix/Cytoperm (BD Biosciences) and stained with V450-conjugated anti-IFNγ (eBioscience). Negative controls without peptide stimulation were run for each sample. All samples were acquired on a BD LSRFortessa (BD Biosciences) flow cytometer and analyzed using FlowJo version 10 software.

### Statistical analysis

PCA analysis was made in R with stats package and visualized with ggplot2 (23) on Vbeta frequencies obtained by flow cytometry.

## Results

### MIS-C presentation overlaps with Toxin Shock Syndrome and KD phenotype

Here, we studied a cohort of 27 children with MISC and compared them with 4 KD diagnosed during the pandemic and 4 retrospective cases of TSS patients. This comparison was motivated by previous descriptions of MIS-C in Europe and in the US that showed a clinical overlap between staphylococcal toxin-mediated TSS and KD in patients with MIS-C. Figure 1 outlines the study flowchart (Figure 1A) and the clinical and biological parameters (Figure 1B) that were evaluated in our study. Children samples were not available for all experiments depending on the volume available. In patients with suspicion of MIS-C, we separated KD from MIS-C based on the positivity of the SARS-CoV2 PCR or serology and the absence of SARS-CoV2 exposure history. All three groups of patients were then subjected to a deep immunological analysis combining cytokine profiling, Vβ analysis and for MIS-C *in vitro* T cell proliferation assays (Figure 1A). We confirmed the strong clinical overlap between MIS-C, TSS and KD. Indeed, many patients in the MIS-C group fulfilled the major criteria for TSS and KD respectively (Figure 1B). Considering the clinical parameters, the most frequent features of MIS-C patients in our cohort were fever, cardiac dysfunction, gastrointestinal symptoms, coagulopathy and inflammation (Table 1).

**Table 1.** Demographic and clinical data of MIS-C patients

| N=27 |  |
| --- | --- |
| **Age, median [Min-Max]** | 9,4 [1,2-15,2] |
| **Female gender** | 8 (30%) |
| **Ethnicity (n, %)** |  |
| Europe | 11 (41%) |
| North Africa | 9 (33%) |
| Subsaharian Africa | 5 (19%) |
| Middle East | 2 (7%) |
| **WHO MIS-C criterias\* (n, %)** |  |
| fever ≥ 3 days | 27 (100%) |
| Rash | 18 (67%) |
| conjunctivitis | 16 (59%) |
| muco-cutaneous inflammation signs<br>(oral, hands or feet) | 16 (59%) |
| Gastrointestinal symptoms<br>(diarrhoea, vomiting, or abdominal<br>pain) | 26 (96%) |
| Hypotension/shock | 11 (41%) |
| **Cardiac dysfunction or abnormalities** |  |
| including elevated Troponin | 19 (70%) |
| including elevated NT-pro-BNP | 23 (85%) |
| **coagulopathy** |  |
| including D-Dimers | 26 (96%) |
| **Inflammatory markers** |  |
| including C-reactive protein | 27 (100%) |
| **Evidence of COVID-19 (n, %)** | 27 (100%) |
| SARS-Cov-2 PCR | 11 (41%) |
| SARS-Cov-2 serology | 24 (89%) |
| **ICU admission** | 19 (70%) |

\* WHO MIS-C criterias ( World Health Organization **Multisystem inflammatory syndrome in children and adolescents with COVID-19: Scientific Brief**, Available at: https://www.who.int/publications-detail/multisystem-inflammatory-syndrome-in-children-and-adolescents-with-covid-19)

**Figure 1:**
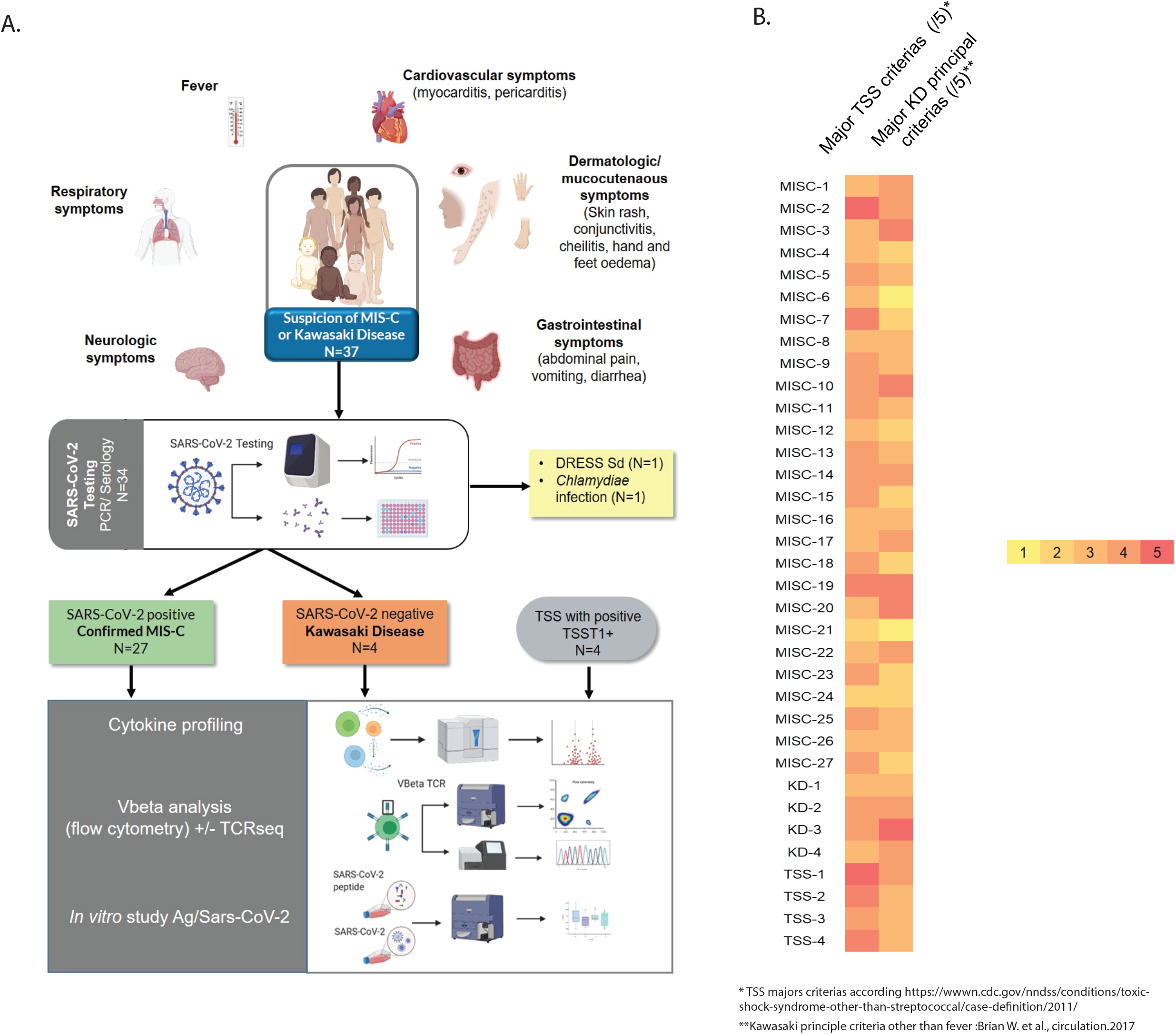
Study design and clinical features of MIS-C patients (A) Outline of the clinical strategy allowing the diagnosis of MIS-C patients and immunological investigation workflow. Patients with symptoms compatible with MIS-C were tested for SARS-CoV2 infection by PCR and serologic tests, which stratified patients in different entities, including SARS-CoV2 negative KD patients. MIS-C, KD and TSS patients were then subjected to different immunological analyses. (B) The heatmap shows the TSS or KD score for each patient calculated as the number of major criteria reached for each disease.

### Strong levels of proinflammatory cytokines in MIS-C contrasting with lymphopenia and low HLA-DR expression in monocytes

SARS-CoV2 can cause fatal acute respiratory distress syndrome in patients at risk. This manifestation is caused by delayed and poorly controlled immune responses, with a deleterious role of inflammatory cytokines. Moreover, we and others have identified a subgroup of severe COVID-19 patients with impaired type-I interferon production(24–27). Thus a regulated production of cytokines is paramount for a good control of SARS-CoV2 infection. This prompted us to investigate how cytokines could contribute to MIS-C pathogenesis. We compared the serum level of IFN-α, IFN-γ, TNF-α, IL-10, soluble CD25 (sCD25), MCP1, IL1Ra and IL-6 between MIS-C, KD, TSS and different forms of COVID-19 (mild pediatric, mild or severe adult-onset COVID-19, Figure 2A-C).

**Figure 2:**
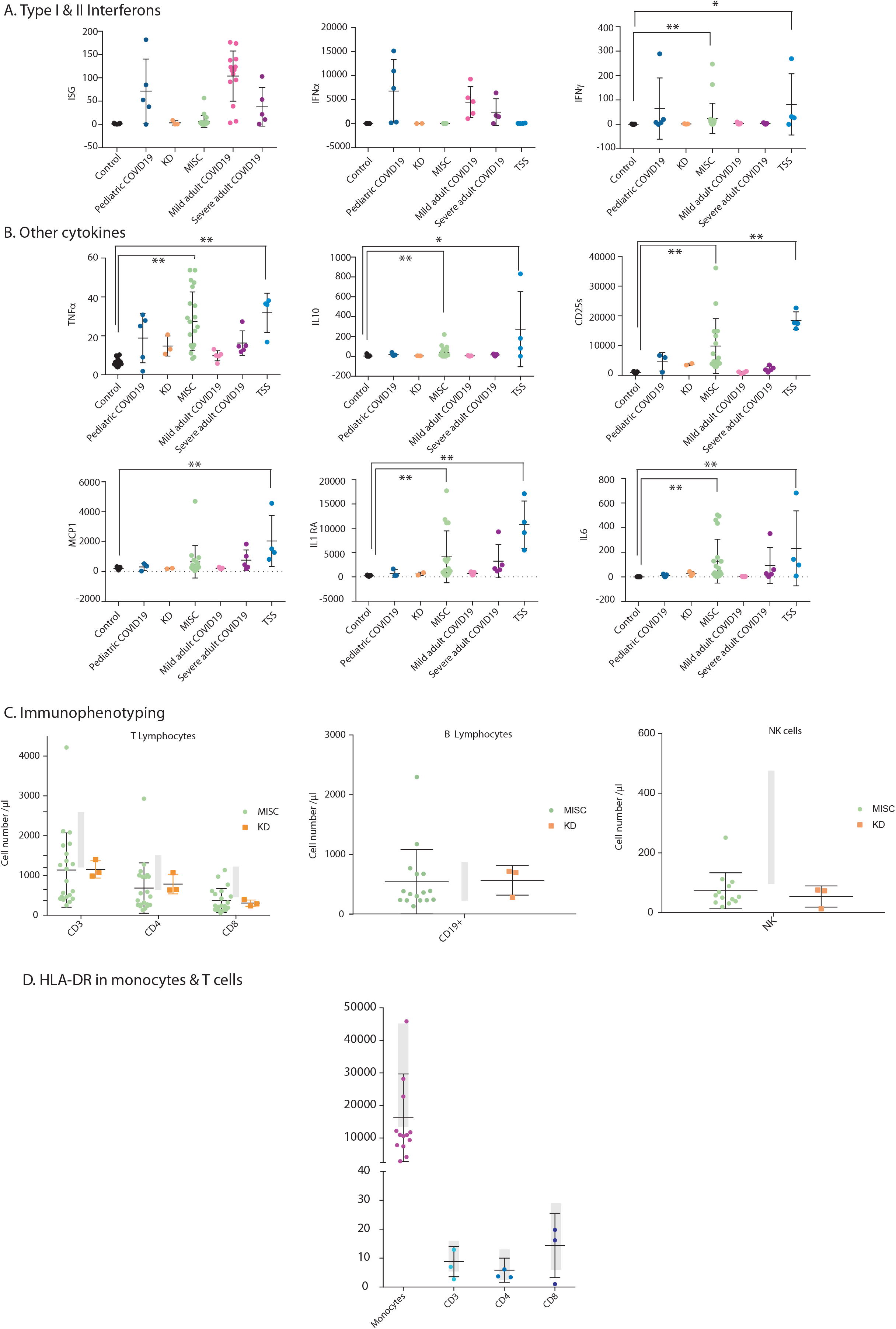
Systemic inflammation and signs of immune paralysis in MIS-C patients (A) Interferon score calculated as the normalized mean expression of six ISGs measured using the Nanostring technology, as previously described(18, 41), Serum IFN-alpha in different groups of patients, as measured with the Simoa technology and IFN-gamma by Elisa (B). Serum levels of the indicated cytokines as measured by automated ELISA. (C) T, B and NK lymphocyte counts measured by flow cytometry in MIS-C and KD (D) HLA-DR expression in T cells and monocytes, as measured by flow cytometry in MIS-C.

Interferons were usually low in MIS-C patients, except for a small subset of patients with high levels of IFN-γ (Figure 2A). The expression of Interferon-stimulated genes (ISG) in blood cells was accordingly lower in MIS-C patients than in patients with different forms COVID-19 (Figure 2B). MIS-C patients were rather characterized by high levels of serum TNF, soluble CD25 (rs IL2), IL-1 Ra, IL-10 and IL-6, which was very similar to TSS patients, and different from COVID-19 patients, irrespective of the form. The few KD patients that we could analyse had rather undetectable levels of serum Interferons (Figure 2A) as well as TNFa, IL10 and sCD25 and detectable but low levels of MCP1, IL6 and IL1RA (Figure 2B). The similarity between MIS-C and TSS cytokines is thus consistent with a cytokine release in MIS-C following superantigenic reaction(28).

To further explore MIS-C immunological profile, we then quantified the number of peripheral lymphocytes of different types, as well as the expression of HLA-DR in patients. T and NK cell counts were on average very low in MISC and KD patients while B cell counts were normal (Figure 2C). HLA-DR is a marker of T and monocyte activation. We found a decreased expression of HLA-DR in monocytes and normal to low values in T cells in both KD and MIS-C patients compared to controls(Figure 2D). This observation is consistent with immune unresponsiveness, as seen following TSS or septic shock(29–32). Indeed, low HLA-DR expression in monocytes is considered as a very good marker of sepsis-induced immunosuppression(32).

### Expansion of Vβ21.3+ peripheral T cells in a large fraction of MIS-C patients

TSST1-related TSS is associated with a skewing of the T cell repertoire towards Vβ2, as a result of TSST1-superantigen induced proliferation of Vβ2+ T cells. Every other S.aureus superantigenic toxins induce a specific TCR Vβ subset, i.e. Vβ 5.2, 5.3, 7.2, 9, 16, 18, 22 for staphylococcal enterotoxin A (SEA) or Vβ 3, 12, 13.2, 14, 17, 20 for SEB(33). The expansion of T cell subsets with a specific Vβ is therefore a strong indication of a superantigenic immune reaction. To explore this phenomenon in MIS-C, we used flow cytometry to assess the distribution of Vβ subunits in T cells from MIS-C patients, in comparison with KD and TSS patients (Figure 3A-C). As expected, TSS patients displayed the hallmark expansion of the Vβ2+ subset in both CD4 and CD8 T cells. Interestingly, several Vβ-specific expansions were also visible in MIS-C patients, and in most cases Vβ21.3+ expansions, in both CD4 and CD8 T subsets. Overall the expansion of Vβ21.3+ T cell subsets was seen in about 40% of MIS-C patients and in none of the few KD patients we could analyse. A principal component analysis of the Vβ distribution in T cells, or in CD4 or CD8 subsets showed that the main parameters separating the different patients were the frequency of Vβ2+ and the frequency of Vβ21.3+ cells (right panels in Figure 3A-C). Thus, the Vβ21.3+ T cell expansion can be considered as a hallmark of MIS-C.

**Figure 3:**
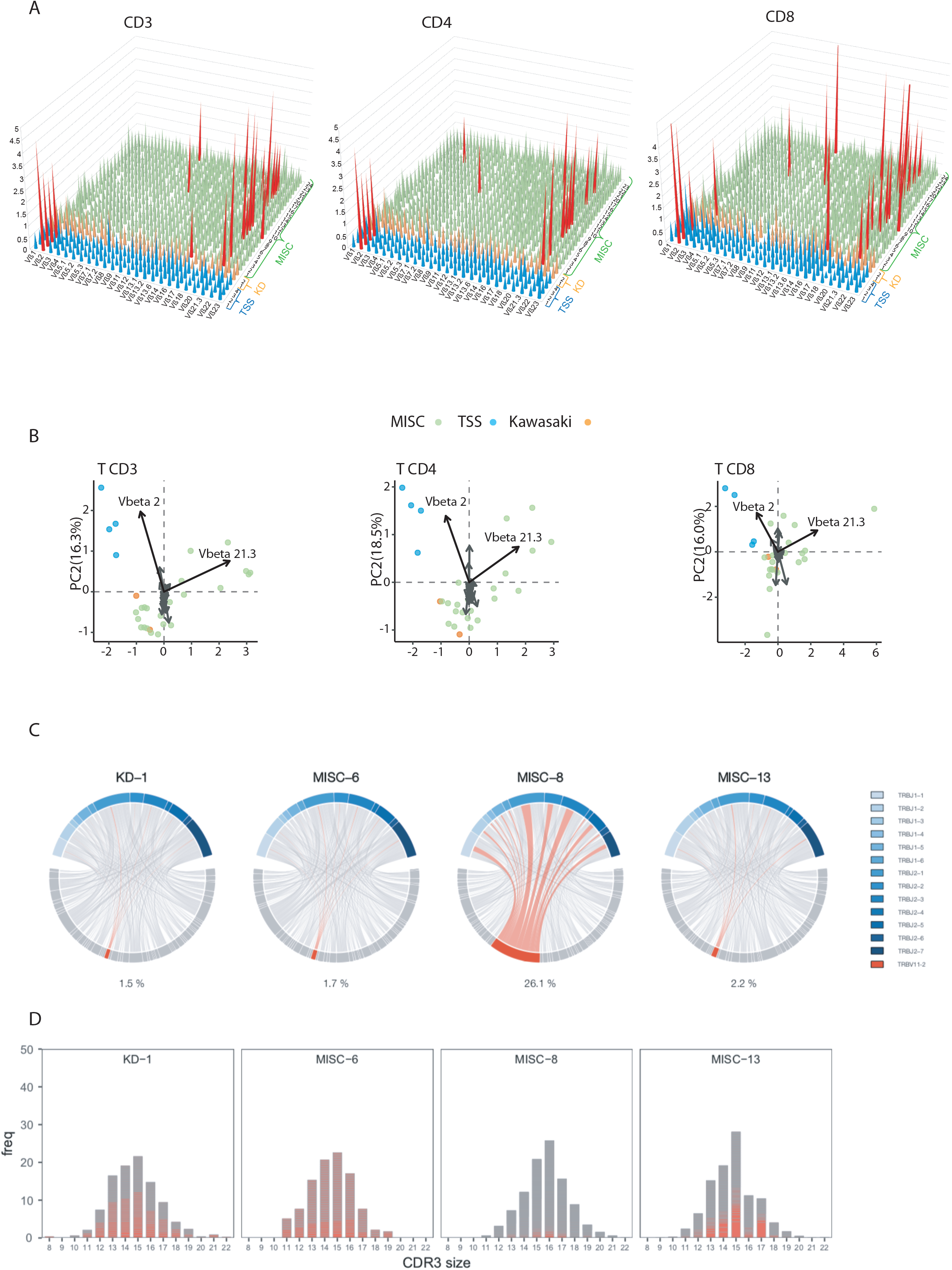
Polyclonal Vb21.3+ T cell expansion in MIS-C patients (A) Frequency of total CD3+ T cells, CD4+ T cells and CD8+ T cells (A) expressing the indicated V-beta (V□) chains, as measured by flow cytometry using specific antibodies against the corresponding Vβ within PBMCs of patients of the indicated group. TSS, KD and MIS-C patients are colored in blue, orange and green respectively. The red color highlights values at least twice higher as the mean frequency in the general adult population. (B) Principal component analysis were made for ach cell population on the basis of the Vb distribution in total, CD4 or CD8 T cells and are shown next to the Vb distributions. Black vectors correspond to the frequency of Vb2+ and Vb21.3+ cells. (C) Chord diagrams of the TRBV (bottom) and TRBJ (top) combinations. Link width is proportional to VJ combination frequency observed in each patient. TRBVs are sorted by alphabetical order, from left to right. TRBJs are color-coded as described in the legend. Combinations using TRBV11-2 are highlighted in red. The percentage values under each chart indicate the percentage of clonotypes expressing TRBV11-2. (D) CDR3 length distribution of clonotypes using TRBV11-2. Each clonotype is represented by a red bar as a function of its CDR3 length in amino-acid. The height of the bar corresponds to the frequency of the clonotype within each repertoire. Grey lines indicate low frequency clonotypes.

### TCR sequencing highlights the polyclonal nature of the TCR V□21.3 expansions

To investigate the clonality of Vβ21.3+ expanded cells, we sequenced and analyzed the TCR repertoires of 4 patients for whom RNA was available (KD-1, MISC-6, MISC-8 and MISC-13). TCRs β chains are generated by recombination of a B gene segment (TRBV) with a J gene segment (TRBJ). TRBV/TRBJ combination usage (Figure 3D) showed the absence of particular expansion in the KD-1, MISC-6, and MISC-13 patients, while 26.1% of MISC-8 circulating T cells used TRBV11-2 (which corresponds to Vβ 21.3 protein). These TRBV11-2 sequences were associated with multiple TRBJ gene, indicating the polyclonal nature of the expansions. This was further confirmed at the level of the complementarity determining region 3 (CDR3), which is the main TCR region that interacts with the cognate antigen. The frequency of TRBV11-2 clonotypes amino-acid sequences as a function of their CDR3 length has a typical gaussian distribution in all patients while clonal expansions should have generated discrete peaks disturbing the gaussian distribution. Besides, we could not identify any specific allele nor mutations of HLA class I or class II genes associated with TRBV11-2 expansions by genomic sequencing of the HLA loci of 7 MIS-C patients (data not shown). Altogether, these results are consistent with a superantigenic origin of the TRBV11-2 expansions in MISC-8.

### Activation of Vβ21.3+ peripheral T cells contrasting with a normal to low SARS-CoV2 peptide specific T cell response

Having shown the polyclonal Vβ21.3+ expansion, we next quested the activation state of this T cell subset. By contrast to the low level of HLA-DR expressed in circulating T cells, we showed a that HLA-DR was massively expressed in the two tested MIS-C patient with Vβ21.3+ expansion while other Vβ (here Vβ1 and Vβ23) were low for HLA-DR expression (Figure 4A), again substantiating the specific activation of Vβ 21.3 in MIS-C, characteristic for a superantigenic immune response. To test the SARS-Cov2 specific T cell activation, we stimulated MIS-C or COVID-19 patient PBMCs with a commercial cocktail of SARS-CoV2 peptides spanning S, N and M viral proteins, reasoning that expanded T cell subsets should not respond to this kind of stimulation if they were induced in a superantigen-like manner. Indeed, T cells from MIS-C patients responded poorly to stimulation with viral peptides, regardless of Vβ21.3 expansion, compared to T cells from COVID-19 patients (Figure 4B). Interestingly, MIS-C T cells also responded poorly to stimulation with a cocktail of EBV/CMV/Influenza-specific peptides, supporting the notion of T cell anergy in MIS-C, a feature also observed in TSS or septic shock (data not shown).

**Figure 4:**
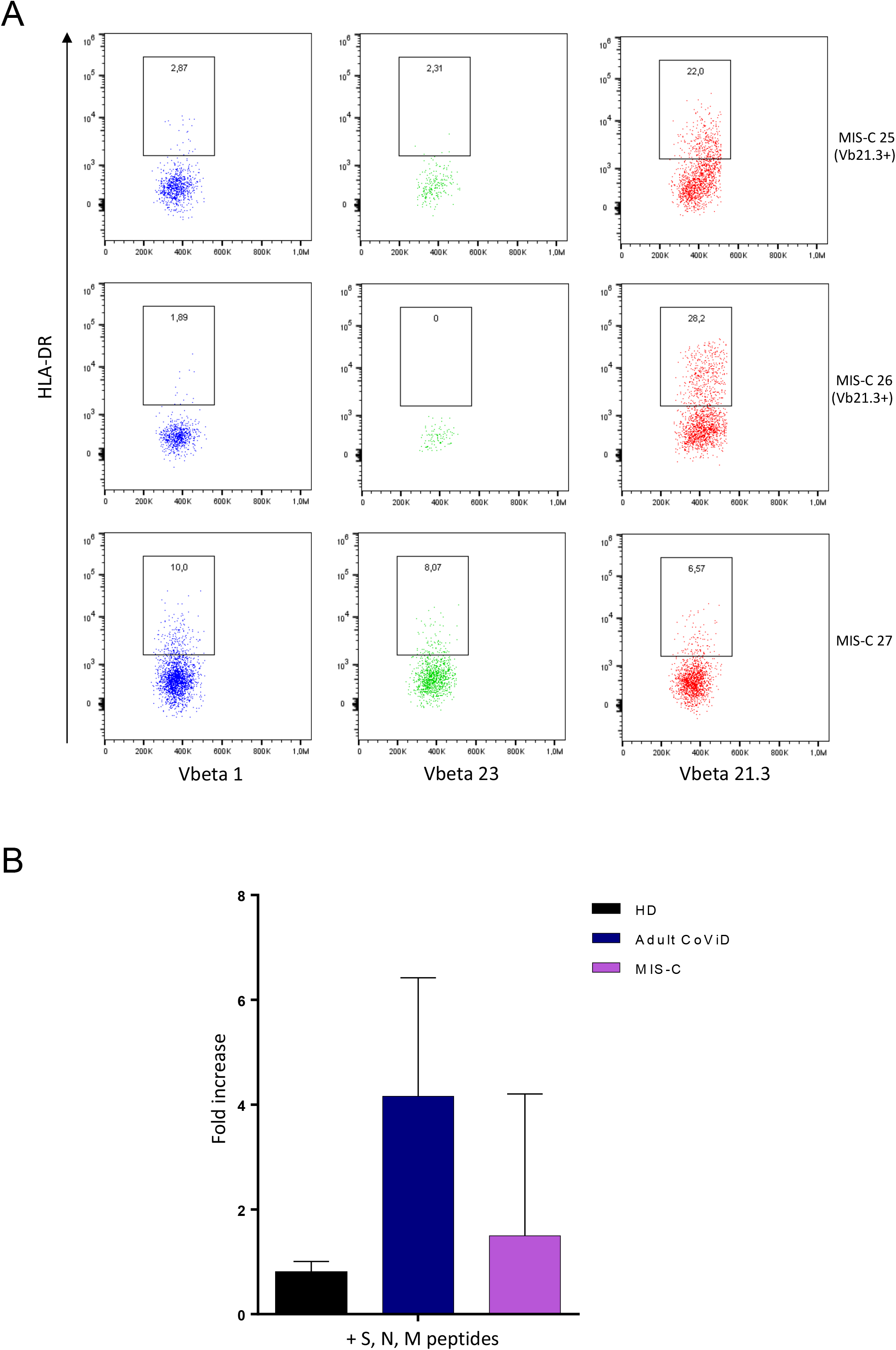
T cell activation within Vβ 21.3 and stimulation of T cells with viral peptides in vitro. (A) Frequency of total T cells, expressing HLA-DR in indicated V-beta (Vβ) chains, as measured by flow cytometry in three MIS-C patients(B) PBMCs from control or MIS-C patients were stimulated for 6h with a commercial cocktail of synthetic peptides from S, N, M SARS-CoV2 peptides in the presence of Golgi secretion inhibitors. Intracellular IFNg expression was then measured in T cells by flow cytometry. N=3 in each group.

Finally, we explored whether the HLA subset could be important in the superantigenic activation of patients, and we further sequenced HLA class I and class II genes in 7 patients and analyzed HLA alleles in 75 additional patients from available exome data by genetic inference but did not identify any specific recurrent subset in MIS-C (data not shown).

## Discussion

The term superantigen (SAg) has been coined by Kappler and Marrack as an operational definition of various T-cell activating substances with specificity for T cell antigen receptors Vβ subunits regardless of the rearrangement and antigen-specificity (34). TSS is a severe toxic shock secondary to the superantigen TSST-1 that stimulates human T cells expressing TCR Vβ2(35). A superantigenic T cell response has been already suspected in KD but the multiple infectious agents associated with this disease hampered the exploration of this phenomenon(14). Yet, many KD features suggest this disease is indeed caused by an infectious agent(36). In particular, the efficacy of IV immunoglobulins, the occurrence in young children, the acute nature of the disease and the absence of relapse suggest that an infectious agent is driving the pathogenesis. The epidemic of a novel coronavirus in 2005 (named New Haven coronavirus) was associated to KD and linked the viral infection to vascular inflammation(37).

Here, we provide compelling evidence showing that MIS-C is caused by a superantigenic T cell reaction. We confirmed the strong overlap in clinical phenotype between MIS-C and TSS, in particular cardiac dysfunction, hypotension, maculo-papular skin rash and conjunctivitis. We also showed a strong similarity in immunological profiles between both clinical entities, in particular a low IFN response but a strong level of different inflammatory cytokines among which IL-6 and TNF. In both diseases, signs of T cell anergy were also visible, such as low expression of HLA-DR and low response to SARS-CoV2 peptides for MIS-C. Most importantly, both TSS and MIS-C are marked by the polyclonal proliferation of a specific Vβ subset i.e. Vβ2+ cells for TSS related to TSST1, and Vβ21.3+ cells for MIS-C. The latter observation was also made in two recent study not yet published, in which patients were sequenced for TCR(38, 39). Porritt et al. discovered an increase of the same TRBV 11-2 (encoding for Vβ21.3 protein subunit of the TCR variable domain)(38) and demonstrated a correlation with disease severity. In addition, using *in silico* modelling, they identified a putative interaction between Vβ21.3 and a superantigen-like motif on the spike of SARS-CoV2. Previous papers have also suggested that the SARS-CoV2 Spike protein could behave as a superantigen structure(40). One of the main challenges now is to understand the cause of the superantigen immune response observed within a window of 3 to 5 weeks after antigen encounter suggesting that both adaptive immune response and the virus itself are necessary to promote aberrant superantigenic immune response(3, 7).

Altogether, our study supports that MIS-C present a superantigenic responses with a specific Vβ21.3 activated T cell expansion and its place for early diagnosis and/or prognosis has to be further explored.

## Data Availability

All data are available upon request

## Author contributions

AB, TW,DK, JM, EM-F designed and analyzed experiments

MM, K LG, AB, CM, RP, SP, SJ, ALM, MP, MV, EC, IR, FV, PB, SYZ performed and analyzed experiments. CM, GM and FV conceptualized the FACS analysis CM and RP supervised cytokine experiments. EJ performed inclusions, chair the clinical investigation and took care of all ethical committee agreement. AP and BK, RP, ST, MG, TL, FV, AMZ, MD, HP, LC, JCR, MM provided clinical samples and clinical details for all cohorts. OT and VB explored HLA in MIS-C patients and JLC, LA supervised genetic inference exploration of HLA. IR and EC provided biobanking and help to generate material for the study TW and AB supervised, designed and funded this study. TW and AB prepared the initial draft. All authors critically reviewed the paper and agreed on the final form.

## Acknowledgments

We acknowledge the patients and families that contributed to this work.

We thank the Square Foundation, Grandir – Fonds de solidarité pour l’enfance.

## References

1. Riphagen S, Gomez X, Gonzalez-Martinez C, Wilkinson N, Theocharis P. Hyperinflammatory shock in children during COVID-19 pandemic. The Lancet 2020;395(10237):1607–1608.

2. Verdoni L et al. An outbreak of severe Kawasaki-like disease at the Italian epicentre of the SARS-CoV-2 epidemic: an observational cohort study [Internet]. The Lancet 2020;0(0). doi:10.1016/S0140-6736(20)31103-X

3. Belot A et al. SARS-CoV-2-related paediatric inflammatory multisystem syndrome, an epidemiological study, France, 1 March to 17 May 2020. Eurosurveillance 2020;25(22):2001010.

4. Levin M. Childhood Multisystem Inflammatory Syndrome - A New Challenge in the Pandemic. N. Engl. J. Med. [published online ahead of print: June 29, 2020]; doi:10.1056/NEJMe2023158

5. Dufort EM et al. Multisystem Inflammatory Syndrome in Children in New York State. N. Engl. J. Med. [published online ahead of print: June 29, 2020]; doi:10.1056/NEJMoa2021756

6. Feldstein LR et al. Multisystem Inflammatory Syndrome in U.S. Children and Adolescents. N. Engl. J. Med. [published online ahead of print: June 29, 2020]; doi:10.1056/NEJMoa2021680

7. Belot A, Levy-Bruhl D, French Covid-19 Pediatric Inflammation Consortium. Multisystem Inflammatory Syndrome in Children in the United States. N Engl J Med 2020;383(18):1793–1794.

8. Consiglio CR et al. The Immunology of Multisystem Inflammatory Syndrome in Children with COVID-19. Cell 2020;183(4):968-981.e7.

9. Diorio C et al. Multisystem inflammatory syndrome in children and COVID-19 are distinct presentations of SARS-CoV-2. J Clin Invest 2020;130(11):5967–5975.

10. Carter MJ et al. Peripheral immunophenotypes in children with multisystem inflammatory syndrome associated with SARS-CoV-2 infection. Nat Med [published online ahead of print: August 18, 2020]; doi:10.1038/s41591-020-1054-6

11. Gruber CN et al. Mapping Systemic Inflammation and Antibody Responses in Multisystem Inflammatory Syndrome in Children (MIS-C). Cell 2020;183(4):982-995.e14.

12. Schvartz A, Belot A, Kone-Paut I. Pediatric Inflammatory Multisystem Syndrome and Rheumatic Diseases During SARS-CoV-2 Pandemic [Internet]. Front. Pediatr. 2020;8. doi:10.3389/fped.2020.605807

13. Casanova J-L, Su HC. A global effort to define the human genetics of protective immunity to SARS-CoV-2 infection [Internet]. Cell 2020;0(0). doi:10.1016/j.cell.2020.05.016

14. Leung DYM, Schlievert PM. Kawasaki syndrome: role of superantigens revisited. FEBS J [published online ahead of print: August 8, 2020]; doi:10.1111/febs.15512

15. Noval Rivas M, Porritt RA, Cheng MH, Bahar I, Arditi M. COVID-19-associated multisystem inflammatory syndrome in children (MIS-C): A novel disease that mimics toxic shock syndrome-the superantigen hypothesis. J Allergy Clin Immunol [published online ahead of print: October 16, 2020]; doi:10.1016/j.jaci.2020.10.008

16. Hoffman M. “Superantigens” may shed light on immune puzzle. Science 1990;248(4956):685–686.

17. Ferry T et al. Analysis of superantigenic toxin Vbeta T-cell signatures produced during cases of staphylococcal toxic shock syndrome and septic shock. Clin Microbiol Infect 2008;14(6):546–554.

18. Pescarmona R et al. Comparison of RT-qPCR and Nanostring in the measurement of blood interferon response for the diagnosis of type I interferonopathies. Cytokine 2019;113:446–452.

19. Gossez M et al. Evaluation of a novel automated volumetric flow cytometer for absolute CD4+ T lymphocyte quantitation. Cytometry B Clin Cytom 2017;92(6):456–464.

20. Demaret J et al. Inter-laboratory assessment of flow cytometric monocyte HLA-DR expression in clinical samples. Cytometry B Clin Cytom 2013;84(1):59–62.

21. Bolotin DA et al. MiXCR: software for comprehensive adaptive immunity profiling. Nature Methods 2015;12(5):380–381.

22. Gu Z, Gu L, Eils R, Schlesner M, Brors B. circlize implements and enhances circular visualization in R. Bioinformatics 2014;30(19):2811–2812.

23. Wickham H. Ggplot2: elegant graphics for data analysis. New York: Springer; 2009:

24. Trouillet-Assant S et al. Type I IFN immunoprofiling in COVID-19 patients.. J Allergy Clin Immunol 2020;146:206-208.e2.

25. Hadjadj J et al. Impaired type I interferon activity and inflammatory responses in severe COVID-19 patients [Internet]. Science [published online ahead of print: July 13, 2020]; doi:10.1126/science.abc6027

26. Bastard P et al. Autoantibodies against type I IFNs in patients with life-threatening COVID-19. Science 2020;370(6515). doi:10.1126/science.abd4585

27. Zhang Q et al. Inborn errors of type I IFN immunity in patients with life-threatening COVID-19. Science 2020;370(6515). doi:10.1126/science.abd4570

28. Faulkner L, Cooper A, Fantino C, Altmann DM, Sriskandan S. The mechanism of superantigen-mediated toxic shock: not a simple Th1 cytokine storm. J Immunol 2005;175(10):6870–6877.

29. Remy S et al. Massive increase in monocyte HLA-DR expression can be used to discriminate between septic shock and hemophagocytic lymphohistiocytosis-induced shock. Crit Care 2018;22(1):213.

30. Monneret G et al. Analytical requirements for measuring monocytic human lymphocyte antigen DR by flow cytometry: application to the monitoring of patients with septic shock. Clin Chem 2002;48(9):1589–1592.

31. The Vβ-specific superantigen staphylococcal enterotoxin B: Stimulation of mature T cells and clonal deletion in neonatal mice. Cell 1989;56(1):27–35.

32. Venet F, Lukaszewicz A-C, Payen D, Hotchkiss R, Monneret G. Monitoring the immune response in sepsis: a rational approach to administration of immunoadjuvant therapies. Curr Opin Immunol 2013;25(4):477–483.

33. Thomas D et al. Staphylococcus aureus superantigens elicit redundant and extensive human Vbeta patterns. Infect Immun 2009;77(5):2043–2050.

34. Kappler J et al. V beta-specific stimulation of human T cells by staphylococcal toxins. Science 1989;244(4906):811–813.

35. Dinges MM, Orwin PM, Schlievert PM. Exotoxins of Staphylococcus aureus. Clinical Microbiology Reviews 2000;13(1):16–34.

36. Leung DY, Meissner C, Fulton D, Schlievert PM. The potential role of bacterial superantigens in the pathogenesis of Kawasaki syndrome. J Clin Immunol 1995;15(6 Suppl):11S–17S.

37. Esper F et al. Association between a Novel Human Coronavirus and Kawasaki Disease. J Infect Dis 2005;191(4):499–502.

38. Porritt RA et al. Identification of a unique TCR repertoire, consistent with a superantigen selection process in Children with Multi-system Inflammatory Syndrome. bioRxiv [published online ahead of print: November 9, 2020]; doi:10.1101/2020.11.09.372169

39. Ramaswamy A et al. Post-infectious inflammatory disease in MIS-C features elevated cytotoxicity signatures and autoreactivity that correlates with severity. medRxiv [published online ahead of print: December 4, 2020]; doi:10.1101/2020.12.01.20241364

40. Cheng MH et al. Superantigenic character of an insert unique to SARS-CoV-2 spike supported by skewed TCR repertoire in patients with hyperinflammation. Proc Natl Acad Sci U S A 2020;117(41):25254–25262.

41. Rice GI et al. Assessment of interferon-related biomarkers in Aicardi-Goutières syndrome associated with mutations in< i> TREX1, RNASEH2A, RNASEH2B, RNASEH2C, SAMHD1</i>, and< i> ADAR</i>: a case-control study. The Lancet Neurology 2013;12(12):1159–1169.

