## Supplemental data for "Superantigenic TCR Vbeta 21.3 signature in Multisystem Inflammatory Syndrome in Children"

Supplemental methods

**Methods**

Patients and ethics

HPI COVID: Written informed consent was obtained for data collection and blood sampling relating to patients and healthy control subjects. The clinical study for children has been registered on ClinicalTrial.gov(NCT04376476) and approved by the national review board for biomedical research in April 2020 (Comité de Protection des Personnes Sud Méditerranée I, Marseille, France) (ID-RCB: 2020-A01102-37).

COVID-SER: For mild adult COVID-19 cohort, the clinical study registered on ClinicalTrial.gov (NCT04341142) has been fully detailed 48. Written informed consent was obtained from all participants and approval was obtained from the national review board for biomedical research in April 2020 (Comité de Protection des Personnes Sud Méditerranée I, Marseille, France; ID RCB 2020-A00932-37).

COVID-rea: For severe adult COVID-19 cohort, this study was registered to the French National Data Protection Agency under the number 20-097 and was approved by an ethical committee for biomedical research (Comité de Protection des Personnes HCL) under the number N°20-41. In agreement with the General Data Protection Regulation (Regulation (EU) 2016/679 and Directive 95/46/EC) and the French data protection law (Law n°78-17 on 06/01/1978 and Décret n°2019-536 on 29/05/2019), we obtained consent from each patient or his next of kin.

T-cell Vβ repertoire analysis

The phenotypic analysis of T-cell Vβ repertoire was performed on whole blood sample using the IOTest Beta Mark kit (Beckman-Coulter) containing 24 monoclonal antibodies (mAbs) identifying ~ 70% of the T cell repertoire. Whole blood cells were stained with APC-Alexa Fluor 750-conjugated anti-CD3, Pacific Blue-conjugated anti-CD4, Krome Orange-conjugated anti-CD8 and each combination of 3 FITC-, PE- and FITC/PE-conjugated anti-Vβ mAbs (Beckman-Coulter) in 8 sample tubes. Whole blood sample were lysed with OptiLyse C Lysing Solution (Beckman-Coulter), washed and fixed in 0.5% formaldehyde in PBS. 0.5 to 10^4^ T cells were acquired on a NAVIOS flow cytometer and data were analysed using NAVIOS software. Lymphocytes were first gated according to FSC/SSC parameter, then by selection of CD3+, CD4+ and CD3+CD4- positive cells. The proportion of each Vβ family was compared to the minimum and the mean+2SD of each reference values obtained from data from IOTest Beta Mark® kit to evaluate expanded or restricted Vβ family. Expansions or restrictions were defined respectively for values above the mean+2SD or below the minimum reference values of the corresponding family.

Lymphocytes immunophenotyping

CD3, CD4 and CD8 T lymphocyte subsets were enumerated on EDTA-anticoagulated peripheral whole blood by single-platform the fully automated volumetric single plateforme technology flow cytometer AQUIOS CL (Beckman-Coulter) as previously described(35).

The phenotypic characterisation of B, NK and T activated lymphocyte subsets were performed on EDTA-anticoagulated whole blood using the following combination of monoclonal antibodies: APC-Alexa Fluor 750-conjugated anti-CD3, Pacific Blue-conjugated anti-CD4, Krome Orange-conjugated anti-CD8, FITC-conjugated anti-HLA-DR, APC-conjugated anti-CD19, Krome Orange-conjugated anti-CD16 and ECD-conjugated anti-CD56 (Beckman-Coulter). The preparations were lysed and fixed by thoroughly mixing and incubating for 10 min successively with 500µL of OptiLyse C reagent (Beckman-Coulter) and 1mL of PBS. The cells were centrifuged for 5min at 400g, resuspended in 500µl of PBS and acquired on a NAVIOS flow cytometer (Beckman-Coulter).
